# Long-read sequencing unravels the complexity of structural variants in *PRKN* in two individuals with early-onset Parkinson’s disease

**DOI:** 10.1101/2024.05.02.24306523

**Authors:** Guillaume Cogan, Kensuke Daida, Kimberley J. Billingsley, Christelle Tesson, Sylvie Forlani, Ludmila Jornea, Lionel Arnaud, Laurene Tissier, Eric LeGuern, Andrew B. Singleton, Mélanie Ferrien, Hélène Gervais Bernard, Suzanne Lesage, Cornelis Blauwendraat, Alexis Brice

**Author notes:** Correspondence to: Alexis Brice, Sorbonne Université, Institut du Cerveau - Paris Brain Institute - ICM, Institut National de la Recherche Médicale-U1127, Centre National de la Recherche Scientifique-UMR7225 Paris France.

## Abstract

**Background:** *PRKN* biallelic pathogenic variants are the most common cause of autosomal recessive early-onset Parkinson’s disease (PD). However, the variants responsible for suspected *PRKN-*PD individuals are not always identified with standard genetic testing.

**Objectives:** Identify the genetic cause in two siblings with a *PRKN*-PD phenotype using long-read sequencing (LRS).

**Methods:** The genetic investigation involved standard testing using successively multiple ligation probe amplification (MLPA), Sanger sequencing, targeted sequencing, whole-exome sequencing and LRS.

**Results:** MLPA and targeted sequencing identified one copy of exon four in *PRKN* but no other variants were identified. Subsequently, LRS unveiled a large deletion encompassing exon 3 to 4 on one allele and a duplication of exon 3 on the second allele; explaining the siblings’ phenotype. MLPA could not identify the balanced rearrangement of exon 3.

**Conclusions:** This study highlights the potential utility of long-read sequencing in the context of unsolved typical *PRKN-*PD individuals.

## Introduction

Parkinson’s disease (PD) is the second most frequent neurodegenerative disorder after Alzheimer’s disease, affecting approximately 6 million people worldwide.^1^ It is characterized by both motor symptoms (bradykinesia, extra-pyramidal rigidity and resting tremor) and non-motor symptoms. Five to 10% of PD cases are monogenic, otherwise PD is generally known to be idiopathic. While more than a dozen genes that contain disease causing mutations have been identified to date, mutations in *PRKN* are most frequently associated with autosomal recessive inheritance and early-onset of disease.^2,3^

*PRKN* spans 1.3 Mb, contains 12 coding exons and is located on chromosome 6q25.2-27.^4^ It encodes for Parkin, a 465 amino-acid E3 ubiquitin-protein ligase, whose main role is maintaining mitochondrial homeostasis.^5^ *PRKN* is located within FRA6E, a known genomic fragile site, subject to structural variations (SVs).^6^ Biallelic pathogenic variants of *PRKN* account for around 4.3% of isolated cases and 8 to 15% of familial young-onset PD ( < 50 years old).^7,8^ The phenotype is usually specific, consisting of a young or juvenile-onset Parkinsonism with a good and longstanding response to levodopa. Dystonia, dyskinesia, and motor fluctuations are typical while autonomic dysfunction, psychotic symptoms, and cognitive decline are usually absent.^9–11^

The recent emergence of long-read sequencing (LRS) has enabled the identification of short tandem repeats and SVs, addressing some of the limitations of short-read sequencing.^12^ The application of LRS in cases of PD where a mutation is suspected, but resistant to identification using traditional methods, could illuminate the genetic cause by overcoming the limitations of short-read sequencing. Here, we describe the detection of complex structural variants by long-read sequencing of two early-onset PD (EOPD) (< 50 years) siblings exhibiting *PRKN-*phenotype left undiagnosed for years after multiple genetic investigations.

## Subjects and Methods

### Study participants

Both participants were extracted based on their early age at onset and their family medical history from a large cohort of 1,587 probands enrolled through the French Parkinson Disease genetics Study Group and international collaborations between 1990 and 2018.^9,13^ PD was diagnosed among the cohort by clinical assessment of movement disorders based on diagnosis criteria from the UK Parkinson Disease Society Brain Bank.

Written informed consent was obtained from participants, and local ethics committees approved genetic studies.

#### Multiplex ligation probe amplification (MLPA), Targeted panel and Whole exome sequencing

MLPA quantifying exons one to 12 of *PRKN* was used to search for rearrangements according to the manufacturer’s instructions (Supplementary methods). A targeted sequencing panel and a whole exome sequencing was performed on individual II-1 as previously described (Supplementary methods).^14,15^ Single Nucleotide Variants (SNVs) were analyzed according to ACMG guidelines and Copy Number Variations (CNVs) to ACMG/AMP guidelines.^16,17^

#### Oxford Nanopore Technologies long-read sequencing and breakpoint region analysis

LRS was performed on II-1’s lymphoblasts sample. Sequencing was prepared according to our protocol reported previously (https://www.protocols.io/view/processing-frozen-cells-for-population-scale-sqk-l-6qpvr347bvmk/v1) (Supplementary methods). We then performed a PCR at the junction breakpoints on both II-1 and II-2 to confirm deletion and duplication events identified with LRS sequences (Supplementary methods, Supplementary Table 1).

#### Data Sharing

Data used in the preparation of this article are available upon request to the authors. Results:

#### Clinical features of the siblings

We report two caucasian siblings presenting with EOPD (Figure 1). Parents were not related and there was no medical history of PD. Both patients presented in their early 30’ with an extrapyramidal syndrome with a good response to levodopa (80-90%), normal cerebral MRI and negative Wilson’s disease biomarkers. The disease slowly evolved and 10 to 15 years after disease-onset they showed no or little nonmotor symptoms. Overall, the clinical features of both individual were highly suggestive of *PRKN-*PD.

**Figure 1.**
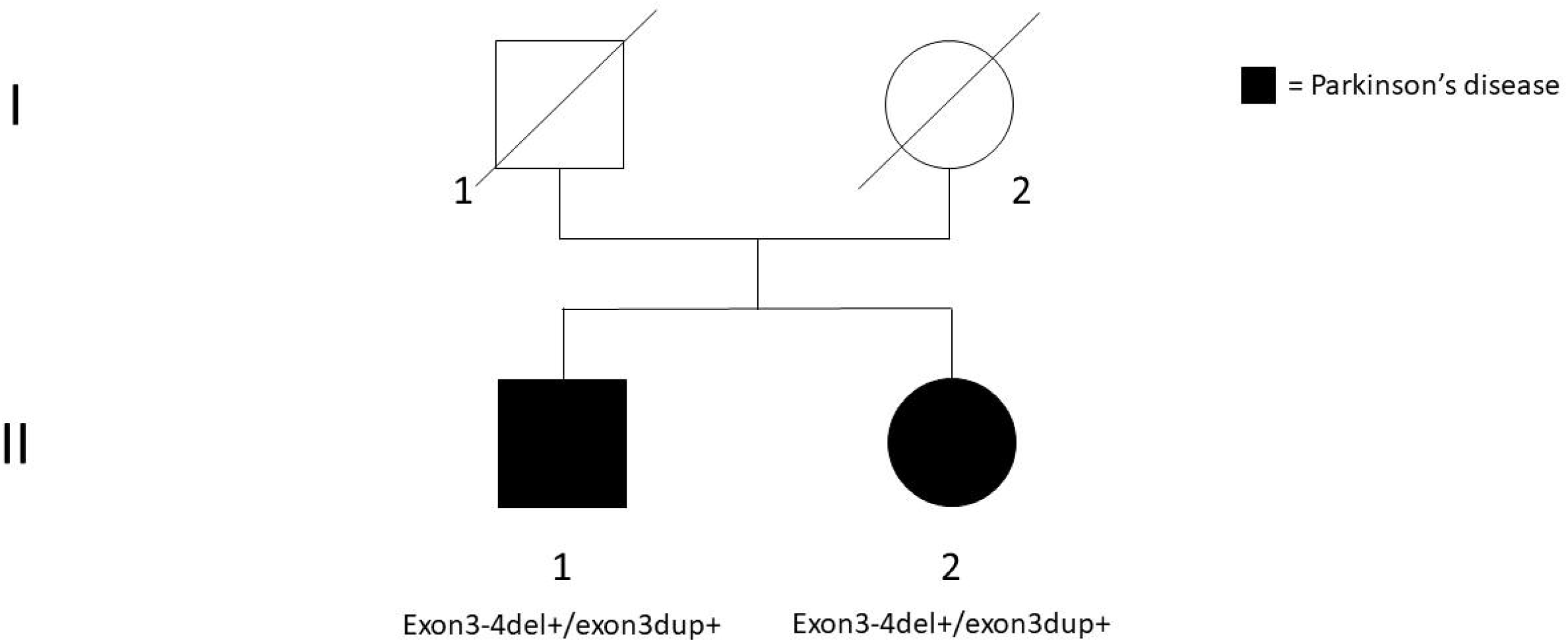
Family pedigree and *PRKN* genotype of the affected siblings (black symbols). The pedigree suggested an autosomal recessive inheritance. Both siblings have two compound heterozygous structural variants: exon 3-4 deletion and exon 3 duplication.

#### Initial genetic investigations

Since the frequency of *PRKN*-PD in early-onset familial PD is relatively high and the disease course was consistent with *PRKN*-PD, we first performed *PRKN* MLPA and Sanger sequencing, which revealed one copy of exon 4 for both individuals and the absence of pathogenic SNV (Figure 2). After the emergence of next generation sequencing, we performed targeted sequencing including *PRKN* on II-1 which confirmed the presence of one copy of exon 4, without any additional pathogenic variant. This result could be interpreted as a heterozygous exon 4 deletion which was not sufficient to explain the phenotype. In addition, WES was performed on II-1 and results were inconclusive with no potential pathogenic variants identified. However, we observed from the raw data that most samples of the run displayed a low coverage of exon 4. Coverage comparison of exon 4 and surrounding exons in *PRKN* revealed a twofold decrease, suggesting that only one copy of exon 4 was present, which was in accordance with previous sequencing methods.

**Figure 2.**
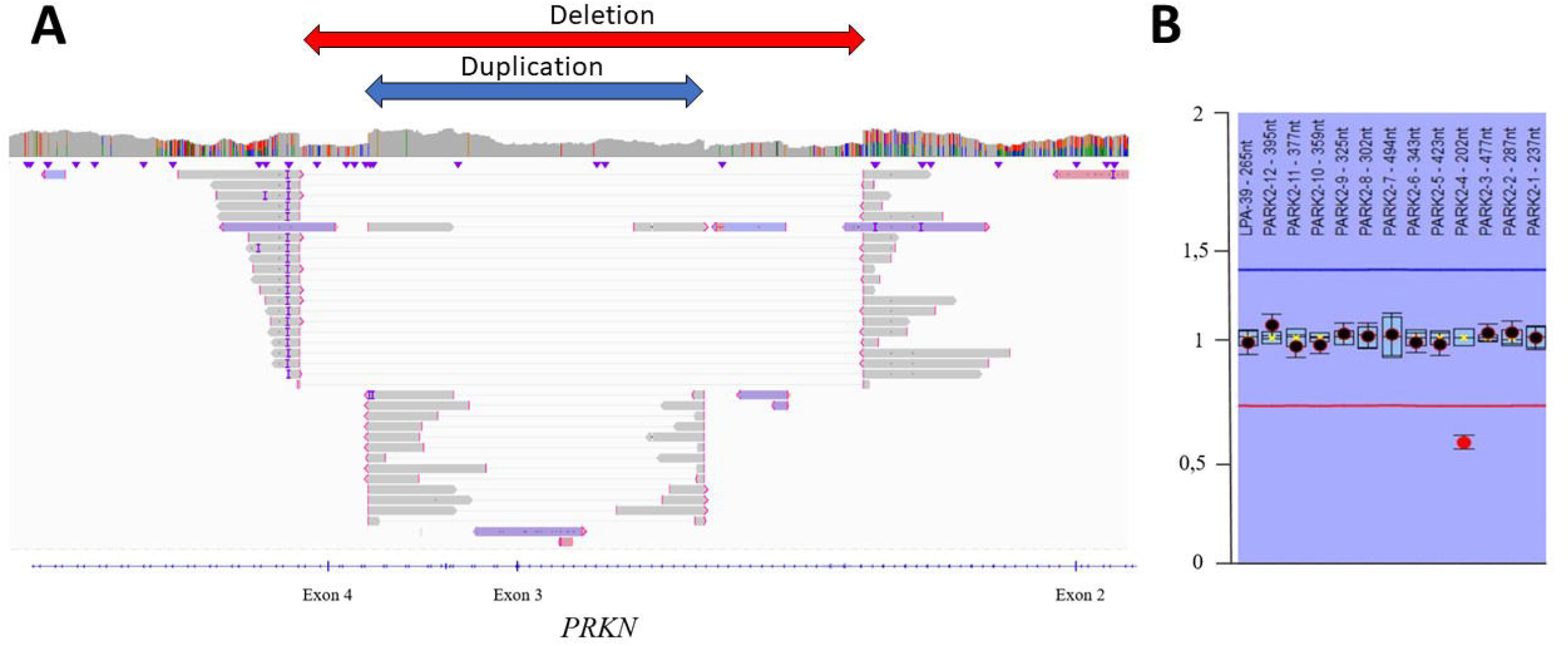
Representation of long-read sequencing (A) and MLPA results (B). (A) Patient II-1 screenshot from Integrative Genome Viewer presenting exon 3-4 deletion (red) and exon 3 duplication in *PRKN* (blue). (B) Exon 3 balanced rearrangement was invisible for MLPA, which only exhibited one copy of exon 4 (ratio: 0.54), suggestive of a heterozygous deletion.

#### Long-read exploration for potential causal variants

We generated LRS data for II-1 using Oxford Nanopore Technologies in addition to the standard methods. Sample DNA QC results were sufficient to perform LRS (Supplementary Figure 1) and overall data output was good with an N50 of 30 and 32.6X estimated coverage (Supplementary table 3).

LRS identified a 106,727 bp (hg38) duplication including exon 3 of *PRKN* (c.(171+1_172-1)_(412+1_413-1)) (Figure 2). Additionally, LRS identified a deletion spanning 178,991 bp (hg38) including exon 3-4 (c.(171+1_172-1)_(534+1_535-1)). Breakpoints are located in deep-intronic regions (supplementary table 2). Both breakpoints of the deletion are located in a short interspersed nuclear element (SINE), either mammalian-wide interspersed repeat (MIR) for intron 2 breakpoint (chr6:162,371,568) or *Alu* for intron 4 breakpoint (chr6:162,192,578). 5’ breakpoint of the duplication was located in a long interspersed nuclear element, whereas the 3’ breakpoint did not fall in any known specific element. Using Integrative Genome Viewer (IGV), we were able to confirm whether these variants were in trans (Figure 2). Exon 3 duplication and exon 3-4 deletions are present in the MDSgene (database), however no coordinates are reported and therefore it is not possible to assess if these are recurring variants. LRS did not identify any additional variants in PD genes (Supplementary table 4). PCR confirmed the presence of the two SVs in II-1 and revealed both variants in II-2 (Supplementary figure 2 and table 3). DNA was not available for other unaffected family members. Altogether, these results demonstrated that biallelic *PRKN* variants were the cause of PD in this family.

## Discussion

*PRKN* is the most frequently mutated gene in autosomal recessive EOPD. However, the genetic cause of patients with a typical *PRKN* phenotype is sometimes elusive because of the limitations of traditional genetic methods to detect more complex structural mutations.^18^

Recently, Daida et al. used LRS to resolve a complex *PRKN* SV in monozygotic twins with PD.^19^ WES and MLPA had first identified a deletion of exon 3. The second variant was a large inversion of 7.4 Mb, not detected by WES and MLPA.

In this study, we identified compound heterozygous SVs in *PRKN* in two siblings with PD. Age of onset and clinical symptoms were similar and suggestive of *PRKN-*PD. One copy of exon 4 was identified in earlier standard genetic investigations. Indeed, the accurate detection of CNV in short-read sequencing is dependent on the ability to detect breakpoints and depth of sequencing. Since both deletion and duplication breakpoints were located in deep intronic regions and genetic dosage of exon 3 was normal, they could not be detected by short-read sequencing. MLPA was consistent with the normal dosage of exon 3, as also shown in a large screening of *PRKN* in our French cohort and solely revealed one copy of exon 4.^9^ Importantly, LRS was able to detect the large 178 kb deletion and 106 kb duplication, encompassing exon 3-4 and exon 3, respectively. Both DNA loss and gain of the same exons 3 and 4 are described in typical *PRKN*-PD individuals as reported in the movement disorders society gene database (https://www.mdsgene.org/d/1/g/4).

Deletions of exon 3 and exon 3-4 of *PRKN* are the most frequent exon rearrangements reported, suggesting recurrent event in intron 2.^20,21^ First, the size of intron 2 is equivalent to that of introns 4, 6, 7, 9 and shorter than intron 1, thereby excluding the hypothesis of a higher frequency due to a size effect. In addition, Mitsui et al. showed that only a minority of breakpoints are recurrent and that various underlying mechanisms are implied in *PRKN* rearrangements.^22^ In contrast to usual techniques, LRS enables the analysis of the breakpoints and gives insights on the mechanism at play. Both deletion breakpoints are located within SINEs which are nonautonomous retrotransposons accounting for 13% of the human genome.^23^ The 5’ and 3’ deletion breakpoints were located in the two different most abundant SINE families: MIR and *Alu*, respectively. The sequence of the breakpoint junction (100 bp upstream and 100 bp downstream) is not associated with either homology nor microhomology. Therefore, a factor affecting replication timing, already known to be implied in PRKN rearrangements, is more likely to explain the deletion than a nonallelic homologous recombination or other mechanisms associated with homo and microhomology.^22^

Standard sequencing techniques are able to identify simple (e.g. coding variants and non complex SVs) biallelic *PRKN* pathogenic variants. However complex *PRKN* SVs, such as balanced rearrangement and inversions, might remain invisible.^9,18^ In addition, patients carrying a single pathogenic variant, as the detected exon 4 deletion in our cases, might have a second hidden causative variant, highlighting the potential need of LRS for *PRKN*-PD diagnosis. In the continuous efforts of studying PD genetics, LRS appears as a useful tool to help identify complex variants (e.g SVs, STRs) and thus novel PD candidate genes. This is exemplified by the LRS identification and recent addition of *RFC1* and *NOTCH2NLC* pathogenic expansions to the known genetic architecture of PD.^8,24^

Since short-read sequencing is unable to solve the genetic cause of many familial and early-onset PD cases, further studies using LRS are required to better decipher the full genetic landscape of PD.

## Data Availability

Data used in the preparation of this article are available upon request to the authors.

## Acknowledgements

We would like to thank all of the participants who donated their time and biological samples to be a part of this study. We also thank Daniella Matute for the help in improving the English quality of the manuscript. Part of this work was carried out the DNA and Cell Bank of the Institut du Cerveau et de la Moëlle épinière (ICM). We gratefully acknowledge Sylvie Forlani and Ludmila Jornea for sample preparation. Part of this work was carried out through the iGenSeq core facility. We gratefully acknowledge Yannick Marie and Agnes Rastetter for the sequencing of targeted panel and whole exome.

This work was supported in part by the Intramural Research Programs of the National Institute on Aging (NIA) and the National Institute of Neurological Disorders and Stroke (NINDS), part of the National Institutes of Health, Department of Health and Human Services (project ZIA AG000949), as well as the Paris Brain Institute and the French Parkinson’s disease genetics study group (PDG). This work utilized the computational resources of the NIH HPC Biowulf cluster (http://hpc.nih.gov).

## Authors’ Roles

Design: G.C, A.S, C.B, A.B

Execution: G.C, K.B, C.T, S.F, L.J, L.A, L.T, E.L, M.F, H.G.B, S.L

Analysis: G.C, K.D, C.T, L.A

Writing: G.C, A.B

Editing of final version: G.C, K.D, L.A, A.S, C.B, A.B

## References

1. GBD 2016 Neurology Collaborators. Global, regional, and national burden of neurological disorders, 1990-2016: a systematic analysis for the Global Burden of Disease Study 2016. Lancet Neurol. 2019;18(5):459–480. doi:10.1016/S1474-4422(18)30499-X

2. Lücking CB, Dürr A, Bonifati V, et al. Association between early-onset Parkinson’s disease and mutations in the parkin gene. N Engl J Med. 2000;342(21):1560–1567. doi:10.1056/NEJM200005253422103

3. Taghavi S, Chaouni R, Tafakhori A, et al. A Clinical and Molecular Genetic Study of 50 Families with Autosomal Recessive Parkinsonism Revealed Known and Novel Gene Mutations. Mol Neurobiol. 2018;55(4):3477–3489. doi:10.1007/s12035-017-0535-1

4. Kitada T, Asakawa S, Hattori N, et al. Mutations in the parkin gene cause autosomal recessive juvenile parkinsonism. Nature. 1998;392(6676):605–608. doi:10.1038/33416

5. Mouton-Liger F, Jacoupy M, Corvol JC, Corti O. PINK1/Parkin-Dependent Mitochondrial Surveillance: From Pleiotropy to Parkinson’s Disease. Front Mol Neurosci. 2017;10:120. doi:10.3389/fnmol.2017.00120

6. Denison SR, Callahan G, Becker NA, Phillips LA, Smith DI. Characterization of FRA6E and its potential role in autosomal recessive juvenile parkinsonism and ovarian cancer. Genes Chromosomes Cancer. 2003;38(1):40–52. doi:10.1002/gcc.10236

7. Kilarski LL, Pearson JP, Newsway V, et al. Systematic review and UK-based study of PARK2 (parkin), PINK1, PARK7 (DJ-1) and LRRK2 in early-onset Parkinson’s disease. Mov Disord Off J Mov Disord Soc. 2012;27(12):1522–1529. doi:10.1002/mds.25132

8. Zhao Y, Qin L, Pan H, et al. The role of genetics in Parkinson’s disease: a large cohort study in Chinese mainland population. Brain J Neurol. 2020;143(7):2220–2234. doi:10.1093/brain/awaa167

9. Lesage S, Lunati A, Houot M, et al. Characterization of Recessive Parkinson Disease in a Large Multicenter Study. Ann Neurol. 2020;88(4):843–850. doi:10.1002/ana.25787

10. Alcalay RN, Caccappolo E, Mejia-Santana H, et al. Cognitive and motor function in long-duration PARKIN-associated Parkinson disease. JAMA Neurol. 2014;71(1):62–67. doi:10.1001/jamaneurol.2013.4498

11. Aasly JO. Long-Term Outcomes of Genetic Parkinson’s Disease. J Mov Disord. 2020;13(2):81–96. doi:10.14802/jmd.19080

12. Method of the Year 2022: long-read sequencing. Nat Methods. 2023;20(1):1. doi:10.1038/s41592-022-01759-x

13. Vollstedt EJ, Kasten M, Klein C, MJFF Global Genetic Parkinson’s Disease Study Group. Using global team science to identify genetic parkinson’s disease worldwide. Ann Neurol. 2019;86(2):153–157. doi:10.1002/ana.25514

14. Casse F, Courtin T, Tesson C, et al. Detection of ATXN2 Expansions in an Exome Dataset: An Underdiagnosed Cause of Parkinsonism. Mov Disord Clin Pract. 2023;10(4):664–669. doi:10.1002/mdc3.13699

15. Bouhouche A, Tesson C, Regragui W, et al. Mutation Analysis of Consanguineous Moroccan Patients with Parkinson’s Disease Combining Microarray and Gene Panel. Front Neurol. 2017;8:567. doi:10.3389/fneur.2017.00567

16. Brandt T, Sack LM, Arjona D, et al. Adapting ACMG/AMP sequence variant classification guidelines for single-gene copy number variants. Genet Med Off J Am Coll Med Genet. 2020;22(2):336–344. doi:10.1038/s41436-019-0655-2

17. Richards S, Aziz N, Bale S, et al. Standards and guidelines for the interpretation of sequence variants: a joint consensus recommendation of the American College of Medical Genetics and Genomics and the Association for Molecular Pathology. Genet Med Off J Am Coll Med Genet. 2015;17(5):405–424. doi:10.1038/gim.2015.30

18. Mor-Shaked H, Paz-Ebstein E, Basal A, et al. Levodopa-responsive dystonia caused by biallelic PRKN exon inversion invisible to exome sequencing. Brain Commun. 2021;3(3):fcab197. doi:10.1093/braincomms/fcab197

19. Daida K, Funayama M, Billingsley KJ, et al. Long-Read Sequencing Resolves a Complex Structural Variant in PRKN Parkinson’s Disease. Mov Disord Off J Mov Disord Soc. 2023;38(12):2249–2257. doi:10.1002/mds.29610

20. Menon PJ, Sambin S, Criniere-Boizet B, et al. Genotype-phenotype correlation in PRKN-associated Parkinson’s disease. NPJ Park Dis. 2024;10(1):72. doi:10.1038/s41531-024-00677-3

21. Hedrich K, Eskelson C, Wilmot B, et al. Distribution, type, and origin of Parkin mutations: review and case studies. Mov Disord Off J Mov Disord Soc. 2004;19(10):1146–1157. doi:10.1002/mds.20234

22. Mitsui J, Takahashi Y, Goto J, et al. Mechanisms of genomic instabilities underlying two common fragile-site-associated loci, PARK2 and DMD, in germ cell and cancer cell lines. Am J Hum Genet. 2010;87(1):75–89. doi:10.1016/j.ajhg.2010.06.006

23. Zhang XO, Pratt H, Weng Z. Investigating the Potential Roles of SINEs in the Human Genome. Annu Rev Genomics Hum Genet. 2021;22:199–218. doi:10.1146/annurev-genom-111620-100736

24. Liu Q, Chen J, Xue J, et al. GGC expansions in NOTCH2NLC contribute to Parkinson disease and dopaminergic neuron degeneration. Eur J Neurol. 2024;31(2):e16145. doi:10.1111/ene.16145

