## Supplementary_material for "Long-read sequencing unravels the complexity of structural variants in *PRKN* in two individuals with early-onset Parkinson’s disease"

**Supplementary Methods.** Multiplex Ligation-dependent Probe Amplification, PCR, Targeted panel, whole exome sequencing, and Oxford Nanopore Technologies long-read sequencing methods

**Supplementary Figure 1.** Gel image for electrophoresis of the PCR products around the deletion and duplication junction points.

**Supplementary Figure 2.** Sample DNA QC results after size selection.

**Supplementary Table 1.** Primer sequence of PCR around junction points.

**Supplementary Table 2.** Location of structural variants breakpoints.

**Supplementary Table 3.** Output data of long-read sequencing.

**Supplementary Table 4.** List of Parkinson’s disease associated genes analyzed by long-read sequencing.

**Supplementary Methods.** Multiplex Ligation-dependent Probe Amplification, PCR, Targeted panel, whole exome sequencing and Oxford Nanopore Technologies long-read sequencing methods

Multiplex ligation probe amplification (MLPA)

Multiplex ligation-dependent probe amplification (MLPA) was used to search *PRKN* rearrangements. MLPA was performed using the MLPA P051-Parkinson mix one kit from MRC-Holland (The Netherlands) according to the manufacturer’s instructions. The kit contains 48 probes, of which 12 are used for quantifying exons one to 12 of the *PRKN* gene. Electrophoresis of the PCR products was performed on an ABI 3730 automated sequencer. MLPA data was analyzed using the Coffalyser software (The Netherlands).

PCR

Junction regions were amplified using PCR with primers designed with Primer3 (Supplementary Table 1). We obtained the full sequence of deleted and duplicated regions using Ensembl (https://www.ensembl.org). A comprehensive view of the breakpoints, design, and validation of the primers was obtained by SnapGene Viewer (<https://www.snapgene.com/snapgene-viewer>). The specificity of the primers was verified using UCSC blat function (<https://genome.ucsc.edu/>). We used RepeatMasker to identify repeating elements surrounding breakpoints and nucleotide BLAST for the analysis of the homology of the breakpoint junction sequences.

Targeted panel

We designed an NGS-based screening of the 22 currently most prevalent parkinsonism-associated genes (Supplementary table 4). The custom Design KAPPA Library Preparation Kit (Roche) was used to capture all exons, intron–exon boundaries, 5′- and 3′-UTR sequences and 10-bp flanking sequences of target genes (RefSeq database, hg19 assembly). Specific probes for NGS target enrichment were designed using NimbleDesign[^1^](https://www.ncbi.nlm.nih.gov/pmc/articles/PMC5674924/#fn1) software and amplicon length varied between 250 and 500 bp. Runs were performed on Illumina MiSeq sequencer. The assay was performed according to the manufacturer’s recommended protocol.

Whole exome sequencing

DNA was extracted from blood of individual II-1. Exons were captured using the Roche Seqcap Ez MedExome (Roche Diagnostics Corporation, Indianapolis, IN) (n = 139) kits followed by 150-bp paired-end sequencing performed on Ilumina NovaSeq 6000 instrument (Illumina Inc, San Diego, CA). Read alignment and variant calling were made using an in-house pipeline. Briefly, FastQC was used to check the quality of the reads and low-quality reads were removed using Trimmomatic. Sequencing data were then aligned to the human reference genome hg19 using the bwa suite and single nucleotide variant (SNV) calling was performed using the HaplotypeCaller function of GATK suite or using Dragen (Illumina). SNV were filtered according to ACMG guidelines : base quality score, location in exons or splice sites, allele frequency (gnomAD). We were also able to detect Copy Number Variants based on WES data generated.  Briefly, the DRAGENTM DNA pipeline v3.8.4 (Illumina) was used to align the reads to the human hg19 reference genome, mark the PCR duplicates and perform the calling of the Copy Number Variants using the panel of normals approach. Within the data set, each sample's depth of coverage being first corrected for the GC bias and then normalized against the depth of all the unrelated samples in the same sequencing batch. Only the events passing the default filters were considered for analysis and annotated with AnnotSV v3.1.1.

Oxford Nanopore technologies long-read sequencing

Briefly, extracted DNA was sized using the Femto Pulse (Agilent Technologies Santa Clara, CA, USA). DNA underwent through a size selection step using the Bluepippin (Sage Science Beverly, MS, USA) to remove fragments below 10 kb. Megaraptor 3 (Diagenode, Belgium) was then used to shear the DNA. Librairies were prepared using the Kit V14 Ligation sequencing kit from ONT and sequenced using PromethION for 72 hours on a R10.4.1 flow cell (Oxford Nanopore Technologies, Oxford, UK). Base calling and mapping to the GRCHh38 reference genome were performed using Guppy v6.3.8 and Winnowmpa v2.0.3, respectively. SNiffles v2.0.7 was used for calling SVs. SVs were annotated by AnnotSV v3.1.1. and CuteSV v1.0.6.

**Supplementary Figure 1.** Gel image for electrophoresis of the PCR product around deletion and duplication junction point.

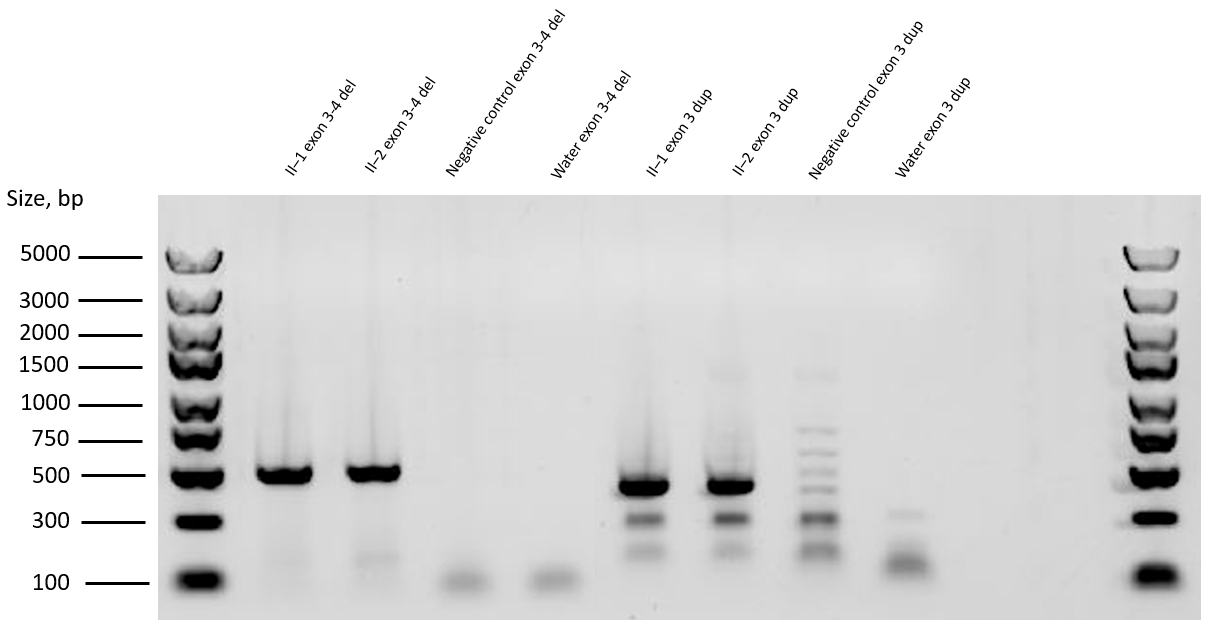

**Supplementary Figure 2.** Individual II–1 sample DNA QC results after size selection.

DNA quality control results after size selection for long-read sequencing.

QC, quality control

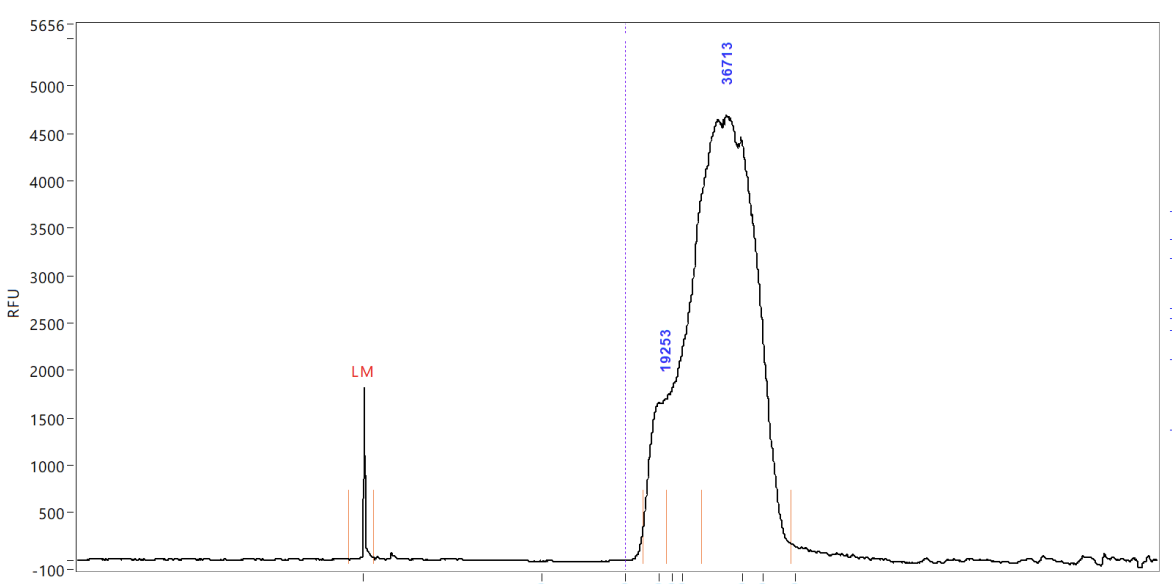

**Supplementary Table 1.** Primer sequence of PCR around junction points.

| Target region | Forward | Reverse |
| --- | --- | --- |
| Deletion junction point | CACGGGCTTTAGTGTCAGGT | TTCACGGCGCTGAGTTTACT |
| Duplication junction point | ACCACGATTCTGCTGCTCAA | GGAAGTTTAGAGGTGGGGCC |

**Supplementary Table 2.** Location of structural variants breakpoints (hg38).

| Structural | 5’ breakpoint | 3’ breakpoint |
| --- | --- | --- |
| Deletion junction point | chr6:162,371,567 - 162,371,568 | chr6:162,192,577 – 162,1692,578 |
| Duplication junction point | chr6:162,321,096 - 162,321,097 | chr6:162,214,370 - 162,214,371 |

**Supplementary Table 3.** Output data of long-read sequencing.

| **SAMPLE** | **Overall output (Gb)** | **Read N50 (kb)** | **Estimated Coverage** |
| --- | --- | --- | --- |
| II-1 | 101.81 | 30.09 | 32.6 |

**Supplementary Table 4.** List of PD genes analyzed by long-read sequencing. Genes sequenced in the targeted panel are highlighted in red.

| ABCA7 | ADH1C | AFG3L2 | ALS2 | AOPEP | AP5Z1 | APOE | APP | ATN1 | ATP13A2 |
| --- | --- | --- | --- | --- | --- | --- | --- | --- | --- |

| ABCA7 | ADH1C | AFG3L2 | ALS2 | AOPEP | AP5Z1 | APOE | APP | ATN1 | ATP13A2 | ATP1A3 | ATP6AP2 | ATP7B | ATXN2 |
| --- | --- | --- | --- | --- | --- | --- | --- | --- | --- | --- | --- | --- | --- |
